# Negative vaccine attitudes and intentions to vaccinate against Covid-19 in relation to smoking status: a population survey of UK adults

**DOI:** 10.1101/2020.12.17.20248396

**Authors:** Sarah E. Jackson, Elise Paul, Jamie Brown, Andrew Steptoe, Daisy Fancourt

## Abstract

**Introduction:** We examined differences in negative attitudes towards vaccines in general, and intentions to vaccinate against Covid-19 specifically, by smoking status in a large sample of adults in the UK.

**Method:** Data were from 29,148 adults participating in the Covid-19 Social Study in September-October 2020. Linear regression analyses examined associations between smoking status (current/former/never) and four types of general negative vaccine attitudes: mistrust of vaccine benefit, worries about unforeseen effects, concerns about commercial profiteering, and preference for natural immunity. Multinomial logistic regression examined associations between smoking status and uncertainty and unwillingness to be vaccinated for Covid-19. Covariates included sociodemographic characteristics and diagnosed health conditions.

**Results:** Relative to never and former smokers, current smokers reported significantly greater mistrust of vaccine benefit, were more worried about unforeseen future effects, had greater concerns about commercial profiteering, and had a stronger preference for natural immunity (*B*_adj_*s* 0.16-0.36, *p*<0.001). Current smokers were more likely to be uncertain (27.6% vs. 22.7% of never smokers: RR_adj_ 1.43 [95%CI 1.31-1.56]; vs. 19.3% of former smokers: RR_adj_ 1.55 [1.41-1.73]) or unwilling (21.5% vs. 11.6% of never smokers: RR_adj_ 2.12 [1.91-2.34]; vs. 14.7% of former smokers: RR_adj_ 1.53 [1.37-1.71]) to receive a Covid-19 vaccine.

**Conclusions:** Current smokers hold more negative attitudes towards vaccines in general, and are more likely to be undecided or unwilling to vaccinate against Covid-19, compared with never and former smokers. With a disproportionately high number of smokers belonging to socially clustered and disadvantaged socioeconomic groups, lower vaccine uptake in this group could also exacerbate health inequalities.

**Implications:** These results suggest that without intervention, smokers will be less likely than non-smokers to take up the offer of a Covid-19 vaccine when offered. Targeted policy action may be required to ensure low uptake of Covid-19 vaccination programmes does not compound health inequalities between smokers and non-smokers.

## Introduction

The Covid-19 pandemic has severely disrupted life in many countries across the world. It is widely accepted that vaccination offers the best chance of a return to normal life, but this relies on adequate uptake to achieve population immunity (1–3). A large body of low certainty evidence suggests current smokers are around 30% less likely than never smokers to become infected with Covid-19 (4,5). The finding has received coverage in social and traditional media. It is possible that smokers may mistakenly interpret this as meaning a vaccine offers little benefit to them and be less likely to take up a vaccine when offered. This could result in smokers being put at more risk than non-smokers and make it more difficult to achieve the vaccine coverage required for population immunity. In addition, by definition, people who smoke are less likely to follow public health advice than those who do not. Smokers tend to have poorer health behaviours than non-smokers, and several studies have indicated that this includes lower uptake of vaccinations (6–8). Understanding whether – and if so, how – attitudes and intentions towards vaccination differ by smoking status is important for informing communications around Covid-19 vaccines. Any disparities could be expressly addressed by correcting misperceptions around smoking and Covid-19 risk – for example, making clear that the risk reduction conferred by vaccination is substantially greater than any uncertain risk reduction associated with smoking or nicotine use.

To our knowledge, no data have been published on the association between smoking status and intentions to vaccinate against Covid-19, although there is evidence of lower uptake of other vaccinations (e.g. influenza) among current smokers than non-smokers (6–8). A comprehensive analysis of negative attitudes towards vaccines and intentions to vaccinate against Covid-19 in relation to sociodemographic and Covid-19 related variables was recently undertaken using data from the Covid-19 Social Study; a large panel study of >70,000 adults in the UK (9). Smoking status was not included in this analysis, but these data are available within the survey. In this study, we use these data to extend current knowledge about anti-vaccine attitudes and intentions to vaccinate against Covid-19 by examining associations with smoking status.

Specifically, we addressed the following research questions:

1. To what extent do negative attitudes towards vaccines in general (mistrust of vaccine benefit, worries about unforeseen future effects, concerns about commercial profiteering, and preference for natural immunity) differ between current, former, and never smokers, adjusting for relevant covariates?
2. To what extent do intentions to vaccinate against Covid-19 – in particular, uncertainty and unwillingness to vaccinate – differ between current, former, and never smokers, adjusting for relevant covariates?

We hypothesised that relative to non-smokers, smokers may (i) have more general mistrust or ambivalence regarding vaccines and (ii) be less likely to intend to vaccinate against Covid-19 (given news reports of a link between smoking and lower risk of Covid-19 infection).

## Method

### Design

We used data from the Covid-19 Social Study. The Covid-19 Social Study is a large panel survey of over 70,000 adults (≥18y) in the UK designed to provide insights into psychological and social experiences during the Covid-19 pandemic. The study commenced on 21 March 2020 and involves online weekly data collection from participants for the duration of the COVID-19 pandemic in the UK.

The study sampling was not random and therefore is not representative of the population, but was intended to have good representation across major sociodemographic groups. The sample has been recruited using three main approaches. First, convenience sampling was used, including promoting the study through existing networks and mailing lists (including large databases of adults who had previously consented to be involved in health research across the UK), social media, and print and digital media coverage. Secondly, more targeted recruitment was undertaken focusing on (i) people from low-income backgrounds, (ii) people with no or few educational qualifications, and (iii) people who were unemployed. Thirdly, the study was promoted via partnerships with third sector organisations to vulnerable groups, including adults with pre-existing mental health conditions, older adults, carers, and people experiencing domestic violence or abuse. The protocol and user guide for the study providing full details on recruitment, retention, and a data dictionary is available on the study website: www.covidsocialstudy.org.

For the present study, we used data collected as part of the vaccine module, which was administered between 7 September and 5 October 2020. We excluded from our analytic sample any participants with missing data on vaccine outcomes, smoking status, or covariates.

### Measures

#### Negative attitudes towards vaccines

Negative general attitudes towards vaccines were measured using the Vaccination Attitudes Examination (VAX) Scale (10). This is a 12-item measure of general attitudes to vaccination with four subscales covering: (i) mistrust of vaccine benefit, (ii) worries about unforeseen future effects, (iii) concerns about commercial profiteering, and (iv) preference for natural immunity. For this study, participants were asked to focus on vaccines in general rather than vaccines for Covid-19 specifically. Each item was rated on a 6-point scale from 1 “strongly agree” to 6 “strongly disagree”, negatively worded items were reverse scored, and scores on items making up each subscale were averaged so that each subscale had a possible score of 1-6. For descriptive purposes, for each subscale those who scored 5 or 6 were considered to have high levels of negative attitudes to vaccines (9).

#### Intention to vaccinate against Covid-19

Uncertainty and unwillingness to vaccinate against Covid-19 when available was assessed with the question: “How likely do you think you are to get a Covid-19 vaccine when one is approved?”. Response options ranged from 1 “very unlikely” to 6 “very likely”. Responses were analysed as an ordinal variable, coded: (0) intend to vaccinate (responses of 5-6), (1) unsure about whether to vaccinate (responses of 3-4), and (2) unwilling to vaccinate (responses of 1-2).

#### Smoking status

Smoking status was assessed with the question: “Do you smoke?” with the response options: (a) non-smoker, (b) ex-smoker, (c) current light smoker (9 or less a day), (d) current moderate smoker (10-19 a day), (e) current heavy smoker (20+ a day). For our analysis, participants answering (c), (d), or (e) were combined as ‘current smokers’.

#### Covariates

We included age, gender, ethnicity, income, key worker status, and diagnosed physical health conditions as covariates in our analysis. Age was analysed as a continuous variable. Gender was categorised as male vs. female (other genders were excluded due to small numbers). Ethnicity was categorised as white vs. ethnic minority groups (i.e. Asian/Asian British, Black/Black British, White and Black/Black British, Mixed race, Chinese/Chinese British, Middle Eastern/Middle Eastern British, or other ethnic group). Those who responded “prefer not to say” on ethnicity were excluded. Annual household income was analysed in five categories: <£16,000, £16,000-29,999, £30,000-59,999, £60,000-89,999, and ≥£90,0000). Key worker status was categorised as key worker vs. not a key worker, with key workers defined as people with jobs deemed by the government to be essential during the pandemic (e.g. health and social care, education and childcare) and who were required to leave home to carry out this work during the lockdown. Participant reports of whether they had received clinical diagnoses of a chronic physical health condition (high blood pressure, diabetes, heart disease, lung disease (asthma or COPD), cancer, or another clinically diagnosed physical health condition) were used to create a binary variable (yes/no) to indicate the presence or absence of pre-existing physical health conditions.

### Statistical analysis

The protocol and analysis plan were pre-registered on Open Science Framework (https://osf.io/xst6z). Analyses were done on complete cases using SPSS v.25. To account for the non-random nature of the sample, all data were weighted to the proportions of gender, age, ethnicity, education, and country of living obtained from the Office for National Statistics (11). We report results on unweighted data in Supplementary File 1 for comparison (there were no notable differences in the pattern of results).

Sample characteristics were summarised using descriptive statistics and compared across current, former, and never smokers using one-way independent analysis of variance and Pearson’s chi-square tests.

Linear regression was used to examine associations between smoking status (never smoker [referent], former smoker, current smoker) and each of the four negative vaccine attitude subscales. We constructed three models: Model 1 was unadjusted, Model 2 was adjusted for sociodemographic characteristics (age, gender, ethnicity, income, and key worker status), and Model 3 was adjusted for sociodemographic characteristics and the presence of ≥1 chronic physical health conditions.

Multinomial logistic regression was used to examine the association between smoking status (never smoker [referent], former smoker, current smoker) and intention to vaccinate against Covid-19. The outcome variable was coded such that (i) uncertainty about whether to vaccinate and (ii) unwillingness to vaccinate were compared against willingness to vaccinate (reference category). We constructed three models with the same adjustments as in the linear regression analysis described above.

In order to assess differences between former and current smokers, we repeated the regression models with former smokers as the referent category.

## Results

A total of 33,082 (unweighted) participants responded to the survey between 7 September and 5 October, of whom 29,148 (unweighted; 28,629 weighted) had complete data and formed the final sample for analysis. Missing cases were primarily due to undisclosed income (*n*=3315), followed by vaccine outcomes (*n*=500), gender (*n*=139), and ethnicity (*n*=98). Table 1 shows sample characteristics in relation to smoking status.

**Table 1.**
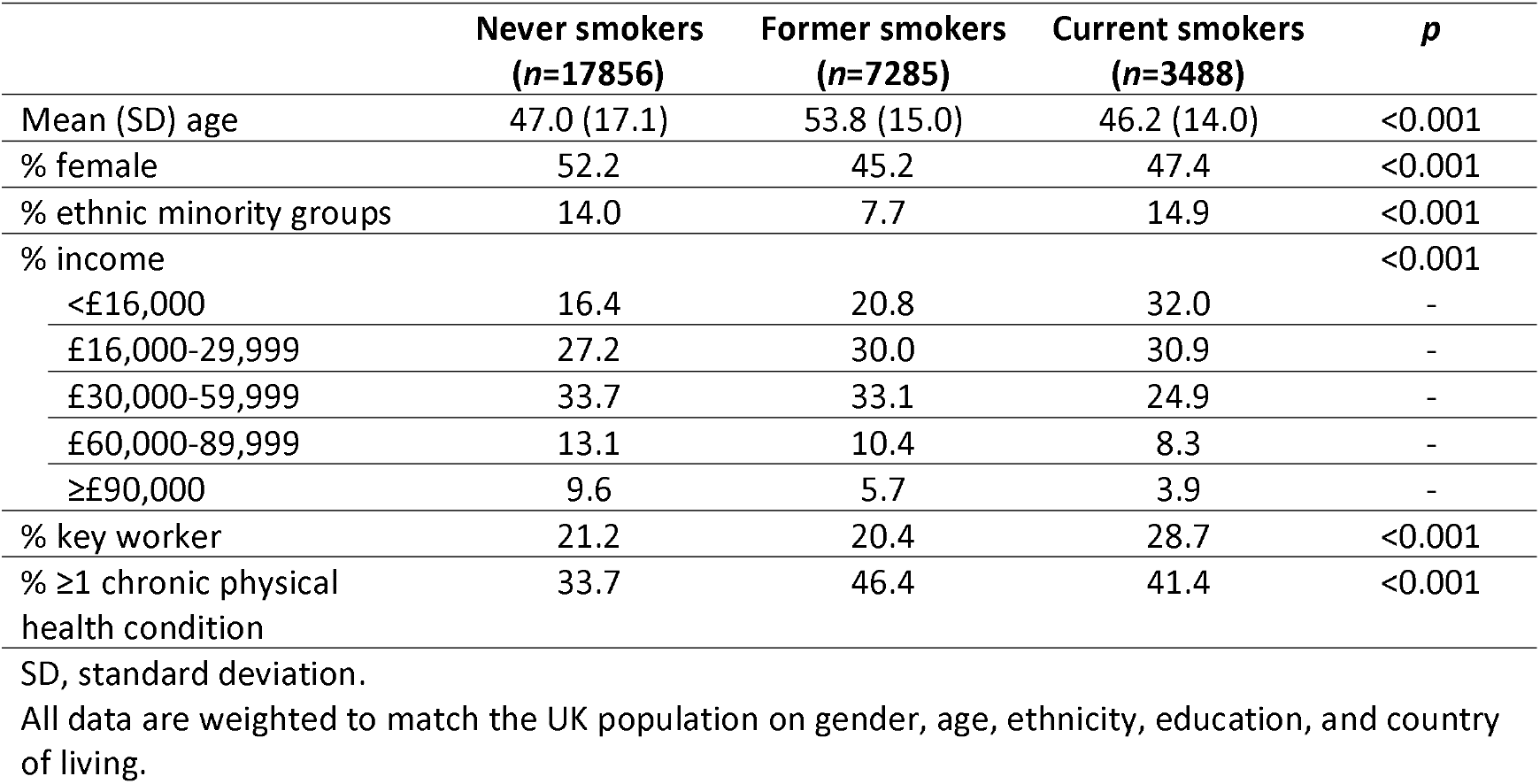
Sample characteristics in relation to smoking status

### Negative vaccine attitudes

The prevalence of high levels of negative attitudes towards vaccines ranged across the four subscales from 5.9-17.7% among never smokers, 8.8-19.4% among former smokers, and 10.6-24.0% among current smokers (Table 2). Relative to never and former smokers, current smokers reported significantly greater mistrust of vaccine benefit, were more worried about unforeseen future effects, had greater concerns about commercial profiteering, and had a stronger preference for natural immunity (Table 2). These differences persisted after adjustment for sociodemographic characteristics and health status. Relative to never smokers, former smokers also scored significantly higher on each domain of negative attitudes towards vaccines, independent of covariates (Table 2).

**Table 2.**
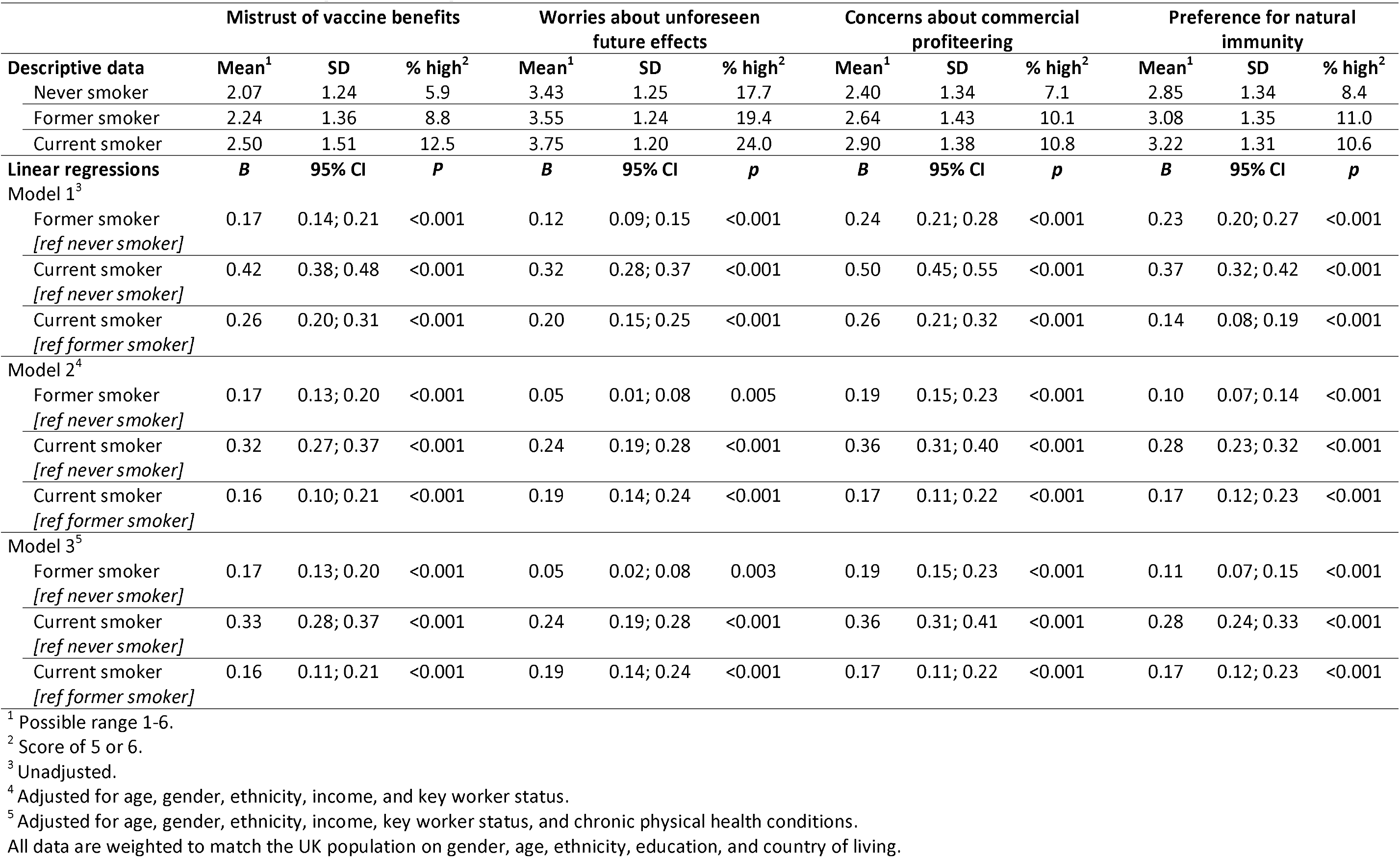
Associations between smoking status and negative attitudes towards vaccines

|  | Mistrust of vaccine benefits |  |  | Worries about unforeseen future effects |  |  | Concerns about commercial profiteering |  |  | Preference for natural immunity |  |  |
| --- | --- | --- | --- | --- | --- | --- | --- | --- | --- | --- | --- | --- |
| Descriptive data | Mean<sup>1</sup> | SD | % high<sup>2</sup> | Mean<sup>1</sup> | SD | % high<sup>2</sup> | Mean<sup>1</sup> | SD | % high<sup>2</sup> | Mean<sup>1</sup> | SD | % high<sup>2</sup> |
| Never smoker | 2.07 | 1.24 | 5.9 | 3.43 | 1.25 | 17.7 | 2.40 | 1.34 | 7.1 | 2.85 | 1.34 | 8.4 |
| Former smoker | 2.24 | 1.36 | 8.8 | 3.55 | 1.24 | 19.4 | 2.64 | 1.43 | 10.1 | 3.08 | 1.35 | 11.0 |
| Current smoker | 2.50 | 1.51 | 12.5 | 3.75 | 1.20 | 24.0 | 2.90 | 1.38 | 10.8 | 3.22 | 1.31 | 10.6 |
| Linear regressions | *B* | 95% CI | *P* | *B* | 95% CI | *p* | *B* | 95% CI | *p* | *B* | 95% CI | *p* |
| Model 1<sup>3</sup> |  |  |  |  |  |  |  |  |  |  |  |  |
| Former smoker<br>[ref never smoker] | 0.17 | 0.14; 0.21 | <0.001 | 0.12 | 0.09; 0.15 | <0.001 | 0.24 | 0.21; 0.28 | <0.001 | 0.23 | 0.20; 0.27 | <0.001 |
| Current smoker<br>[ref never smoker] | 0.42 | 0.38; 0.48 | <0.001 | 0.32 | 0.28; 0.37 | <0.001 | 0.50 | 0.45; 0.55 | <0.001 | 0.37 | 0.32; 0.42 | <0.001 |
| Current smoker<br>[ref former smoker] | 0.26 | 0.20; 0.31 | <0.001 | 0.20 | 0.15; 0.25 | <0.001 | 0.26 | 0.21; 0.32 | <0.001 | 0.14 | 0.08; 0.19 | <0.001 |
| Model 2<sup>4</sup> |  |  |  |  |  |  |  |  |  |  |  |  |
| Former smoker<br>[ref never smoker] | 0.17 | 0.13; 0.20 | <0.001 | 0.05 | 0.01; 0.08 | 0.005 | 0.19 | 0.15; 0.23 | <0.001 | 0.10 | 0.07; 0.14 | <0.001 |
| Current smoker<br>[ref never smoker] | 0.32 | 0.27; 0.37 | <0.001 | 0.24 | 0.19; 0.28 | <0.001 | 0.36 | 0.31; 0.40 | <0.001 | 0.28 | 0.23; 0.32 | <0.001 |
| Current smoker<br>[ref former smoker] | 0.16 | 0.10; 0.21 | <0.001 | 0.19 | 0.14; 0.24 | <0.001 | 0.17 | 0.11; 0.22 | <0.001 | 0.17 | 0.12; 0.23 | <0.001 |
| Model 3<sup>5</sup> |  |  |  |  |  |  |  |  |  |  |  |  |
| Former smoker<br>[ref never smoker] | 0.17 | 0.13; 0.20 | <0.001 | 0.05 | 0.02; 0.08 | 0.003 | 0.19 | 0.15; 0.23 | <0.001 | 0.11 | 0.07; 0.15 | <0.001 |
| Current smoker<br>[ref never smoker] | 0.33 | 0.28; 0.37 | <0.001 | 0.24 | 0.19; 0.28 | <0.001 | 0.36 | 0.31; 0.41 | <0.001 | 0.28 | 0.24; 0.33 | <0.001 |
| Current smoker<br>[ref former smoker] | 0.16 | 0.11; 0.21 | <0.001 | 0.19 | 0.14; 0.24 | <0.001 | 0.17 | 0.11; 0.22 | <0.001 | 0.17 | 0.12; 0.23 | <0.001 |

<sup>1</sup> Possible range 1-6.<sup>2</sup> Score of 5 or 6.<sup>3</sup> Unadjusted.<sup>4</sup> Adjusted for age, gender, ethnicity, income, and key worker status.<sup>5</sup> Adjusted for age, gender, ethnicity, income, key worker status, and chronic physical health conditions.
All data are weighted to match the UK population on gender, age, ethnicity, education, and country of living.

### Intention to vaccinate against Covid-19

65.8% (95% CI 65.1-66.4%) of never smokers, 66.0% (64.9-67.1%) of former smokers, and 50.9% (49.3-52.6%) of current smokers said they intend to receive the Covid-19 vaccine when one becomes available. There was a graded association between smoking status and lack of intent to vaccinate, with never smokers the least likely and current smokers most likely to report being unwilling to vaccinate against Covid-19 (Table 3). In addition, relative to never smokers, former smokers were less likely and current smokers were more likely to report being uncertain (Table 3). These differences persisted after adjustment for sociodemographic characteristics and health status.

**Table 3.** Associations between smoking status and uncertainty and unwillingness to vaccinate against Covid-19

|  | Undecided |  |  | Unwilling |  |  |
| --- | --- | --- | --- | --- | --- | --- |
| **Descriptive data** | **%** | **95% CI** | **-** | **%** | **95% CI** | **-** |
| Never smoker | 22.7 | 22.1; 23.3 | - | 11.6 | 11.1; 12.1 | - |
| Former smoker | 19.3 | 18.4; 20.2 | - | 14.7 | 13.9; 15.5 | - |
| Current smoker | 27.6 | 26.1; 29.1 | - | 21.5 | 20.2; 22.9 | - |
| **Multinomial logistic regressions** | **RR** | **95% CI** | **p** | **RR** | **95% CI** | **p** |
| **Model 1<sup>1</sup>** |  |  |  |  |  |  |
| Former smoker<br>*[ref never smoker]* | 0.85 | 0.79; 0.91 | <0.001 | 1.26 | 1.17; 1.37 | 0.067 |
| Current smoker<br>*[ref never smoker]* | 1.57 | 1.44; 1.71 | <0.001 | 2.40 | 2.18; 2.64 | <0.001 |
| Current smoker<br>*[ref former smoker]* | 1.85 | 1.68; 2.04 | <0.001 | 1.90 | 1.70; 2.11 | <0.001 |
| **Model 2<sup>2</sup>** |  |  |  |  |  |  |
| Former smoker<br>*[ref never smoker]* | 0.89 | 0.83; 0.96 | 0.002 | 1.35 | 1.24; 1.47 | <0.001 |
| Current smoker<br>*[ref never smoker]* | 1.39 | 1.27; 1.52 | <0.001 | 2.04 | 1.85; 2.26 | <0.001 |
| Current smoker<br>*[ref former smoker]* | 1.55 | 1.40; 1.72 | <0.001 | 1.52 | 1.36; 1.70 | <0.001 |
| **Model 3<sup>3</sup>** |  |  |  |  |  |  |
| Former smoker<br>*[ref never smoker]* | 0.92 | 0.85; 0.98 | 0.015 | 1.39 | 1.28; 1.51 | <0.001 |
| Current smoker<br>*[ref never smoker]* | 1.43 | 1.31; 1.56 | <0.001 | 2.12 | 1.91; 2.34 | <0.001 |
| Current smoker<br>*[ref former smoker]* | 1.56 | 1.41; 1.73 | <0.001 | 1.53 | 1.37; 1.71 | <0.001 |

<sup>1</sup> Unadjusted.
<sup>2</sup> Adjusted for age, gender, ethnicity, income, and key worker status.
<sup>3</sup> Adjusted for age, gender, ethnicity, income, key worker status, and chronic physical health conditions.
All data are weighted to match the UK population on gender, age, ethnicity, education, and country of living.

## Discussion

In a large sample of adults in the UK, we documented notable associations between smoking status and attitudes towards vaccination against Covid-19. In general (i.e. not specific to Covid-19), current smokers reported the greatest levels (and never smokers the lowest levels) of mistrust in the benefit of vaccines, worries about unforeseen future effects, concerns about commercial profiteering, and preference for natural immunity. These attitudes have been shown to be strongly associated with uptake of other vaccines (e.g. influenza) (12). When asked whether they would take up the offer of a Covid-19 vaccination when one becomes available, current smokers were the most likely to report being uncertain or unwilling, with just 51% reporting an intention to vaccinate compared with 66% of former smokers and never smokers. These differences were independent of age, gender, ethnicity, income, key worker status, or chronic physical health conditions.

These findings are in line with previous research documenting lower uptake of vaccination for other viruses (e.g. influenza) among smokers than non-smokers (6,7). It is not clear in this case whether differences are attributable to smokers being aware of the link between smoking and lower risk of Covid-19 infection (4,5), or are the product of a more general mistrust of vaccines or propensity to engage in health-risk behaviours. In either case, these results have important implications for public health. Without intervention, smokers appear less likely to take up the offer of a Covid-19 vaccine when one is available. This could impede efforts to achieve the level of population immunity required to facilitate a return to unrestricted living. With a disproportionately high number of smokers belonging to disadvantaged socioeconomic groups (13) – which have been hit harder by the pandemic (14) – lower vaccine uptake in this group could also exacerbate health inequalities. Smoking also tends to cluster strongly in social networks (15) and groups with low vaccine uptake who socialise together are likely to continue to be disproportionately affected by Covid-19.

There is potential to address the vaccination intention gap through targeted messaging and support aimed at smokers. Over a quarter of current smokers in our sample reported being undecided about whether to vaccinate against Covid-19. There will be a range of options available to policymakers to try and change the behaviour of priority subgroups towards receiving a Covid-19 vaccine (16). One option may be to target increasing smokers’ awareness of the benefits of vaccination and correcting any potential misperceptions around smoking conferring protection against Covid-19 could encourage those who are undecided – and perhaps even those who are currently unwilling – to accept a vaccine when offered (17). Other policies options may include those related to incentivisation, modelling, environmental restructuring or enablement.

Strengths of this study include the large sample size and collection of data in real-time during the second wave of the Covid-19 pandemic in the UK, just prior to positive vaccine trial results being reported and the first vaccine being approved for use in the UK. A key limitation is the non-random sampling method, although we corrected for this by weighting the data to match the population and observed no notable differences between weighted and unweighted results.

In conclusion, in the UK, current and former smokers hold more negative attitudes than never smokers towards vaccines in general, and may be less likely to accept a Covid-19 vaccination when offered. Targeted policy action may be required to ensure low uptake of Covid-19 vaccination programmes does not compound inequalities in health between smokers and non-smokers.

## Data Availability

The COVID-19 Social Study documentation and codebook are available for download at https://www.covidsocialstudy.org/.

## Declarations

### Ethics approval and consent to participate

Ethical approval for the COVID-19 Social Study was granted by the UCL Ethics Committee [12467/005]. All participants provided fully informed consent and the study is GDPR compliant.

## References

1. Shin MD, Shukla S, Chung YH, Beiss V, Chan SK, Ortega-Rivera OA, et al. COVID-19 vaccine development and a potential nanomaterial path forward. Nat Nanotechnol. 2020 Aug;15(8):646–55.

2. Jeyanathan M, Afkhami S, Smaill F, Miller MS, Lichty BD, Xing Z. Immunological considerations for COVID-19 vaccine strategies. Nat Rev Immunol. 2020 Oct;20(10):615–32.

3. World Health Organization. Draft landscape of COVID-19 candidate vaccines [Internet]. 2020 [cited 2020 Nov 18]. Available from: https://www.who.int/publications/m/item/draft-landscape-of-covid-19-candidate-vaccines

4. Simons D, Shahab L, Brown J, Perski O. The association of smoking status with SARS-CoV-2 infection, hospitalisation and mortality from COVID-19: A living rapid evidence review with Bayesian meta-analyses (version 9). Qeios [Internet]. 2020 Nov 6 [cited 2020 Nov 17]; Available from: https://www.qeios.com/read/UJR2AW.10

5. Simons D, Shahab L, Brown J, Perski O. The association of smoking status with SARS-CoV-2 infection, hospitalisation and mortality from COVID-19: A living rapid evidence review with Bayesian meta-analyses (version 7). Addiction [Internet]. [cited 2020 Nov 17];n/a(n/a). Available from: https://onlinelibrary.wiley.com/doi/abs/10.1111/add.15276

6. Wada K, Smith DR. Influenza Vaccination Uptake among the Working Age Population of Japan: Results from a National Cross-Sectional Survey. PLOS ONE. 2013 Mar 12;8(3):e59272.

7. Mangtani P, Breeze E, Kovats S, Ng ESW, Roberts JA, Fletcher A. Inequalities in influenza vaccine uptake among people aged over 74 years in Britain. Prev Med. 2005 Aug 1;41(2):545–53.

8. Lu P, Gonzalez-Feliciano A, Ding H, Bryan LN, Yankey D, Monsell EA, et al. Influenza A (H1N1) 2009 monovalent and seasonal influenza vaccination among adults 25 to 64 years of age with high-risk conditions--United States, 2010. Am J Infect Control. 2013 Aug;41(8):702–9.

9. Paul E, Steptoe A, Fancourt D. Attitudes towards vaccines and intention to vaccinate against COVID-19: Implications for public health communications. Lancet Reg Health - Eur. in press;

10. Martin LR, Petrie KJ. Understanding the Dimensions of Anti-Vaccination Attitudes: the Vaccination Attitudes Examination (VAX) Scale. Ann Behav Med. 2017 Oct 1;51(5):652–60.

11. Office for National Statistics. Population estimates [Internet]. 2018 [cited 2020 Apr 25]. Available from: https://www.ons.gov.uk/peoplepopulationandcommunity/populationandmigration/populationestimates

12. Nicholls LAB, Gallant A, Cogan N, Rasmussen S, Young D, Williams L. Older adults’ vaccine hesitancy: psychosocial factors associated with influenza, pneumococcal, and shingles vaccine uptake. [Internet]. PsyArXiv; 2020 Oct [cited 2020 Dec 15]. Available from: https://psyarxiv.com/xzb8y/

13. Hiscock R, Bauld L, Amos A, Fidler JA, Munafò M. Socioeconomic status and smoking: a review. Ann N Y Acad Sci. 2012 Feb;1248:107–23.

14. Patel JA, Nielsen FBH, Badiani AA, Assi S, Unadkat VA, Patel B, et al. Poverty, inequality and COVID-19: the forgotten vulnerable. Public Health. 2020 Jun;183:110–1.

15. Christakis NA, Fowler JH. The collective dynamics of smoking in a large social network. N Engl J Med. 2008 May 22;358(21):2249–58.

16. Michie S, van Stralen MM, West R. The behaviour change wheel: A new method for characterising and designing behaviour change interventions. Implement Sci. 2011 Apr 23;6:42.

17. World Health Organization. Behavioural considerations for acceptance and uptake of COVID-19 vaccines [Internet]. 2020 [cited 2020 Dec 15]. (WHO technical advisory group on behavioural insights and sciences for health, meeting report, 15 October 2020). Available from: https://www.who.int/publications-detail-redirect/9789240016927

