## Supplementary File 1 for "Negative vaccine attitudes and intentions to vaccinate against Covid-19 in relation to smoking status: a population survey of UK adults"

| **Supplementary Table 1.** Sample characteristics in relation to smoking status: unweighted data | | | | | |
| --- | --- | --- | --- | --- | --- |
|  | | **Never smokers**  **(*n*=19586)** | **Former smokers**  **(*n*=7330)** | **Current smokers**  **(*n*=2232)** | ***p*** |
| Mean (SD) age | | 53.2 (14.5) | 56.8 (13.0) | 51.5 (12.5) | <0.001 |
| % female | | 75.7 | 70.4 | 74.3 | <0.001 |
| % ethnic minority groups | | 3.9 | 2.6 | 4.7 | <0.001 |
| % income | |  |  |  | <0.001 |
|  | <£16,000 | 12.3 | 17.8 | 28.0 | - |
|  | £16,000-29,999 | 24.2 | 28.1 | 29.5 | - |
|  | £30,000-59,999 | 36.1 | 33.8 | 28.5 | - |
|  | £60,000-89,999 | 16.1 | 12.5 | 8.6 | - |
|  | ≥£90,000 | 11.3 | 7.9 | 5.3 | - |
| % key worker | | 21.2 | 20.1 | 26.3 | <0.001 |
| % ≥1 chronic physical health condition | | 36.7 | 47.4 | 42.5 | <0.001 |
| SD, standard deviation. | | | | | |

| **Supplementary Table 2.** Associations between smoking status and negative attitudes towards vaccines: unweighted data | | | | | | | | | | | | | | | | |
| --- | --- | --- | --- | --- | --- | --- | --- | --- | --- | --- | --- | --- | --- | --- | --- | --- |
|  | | **Mistrust of vaccine benefits** | | |  | **Worries about unforeseen future effects** | | |  | **Concerns about commercial profiteering** | | |  | **Preference for natural immunity** | | |
| **Descriptive data** | | **Mean^1^** | **SD** | **% high^2^** |  | **Mean^1^** | **SD** | **% high^2^** |  | **Mean^1^** | **SD** | **% high^2^** |  | **Mean^1^** | **SD** | **% high^2^** |
|  | Never smoker | 1.96 | 1.16 | 4.8 |  | 3.35 | 1.23 | 15.3 |  | 2.19 | 1.26 | 5.2 |  | 2.72 | 1.30 | 6.5 |
|  | Former smoker | 2.07 | 1.22 | 5.9 |  | 3.45 | 1.24 | 17.4 |  | 2.38 | 1.34 | 6.8 |  | 2.86 | 1.32 | 7.7 |
|  | Current smoker | 2.28 | 1.38 | 8.6 |  | 3.67 | 1.25 | 22.9 |  | 2.73 | 1.38 | 9.3 |  | 3.09 | 1.33 | 10.0 |
| **Linear regressions** | | ***B*** | **95% CI** | ***p*** |  | ***B*** | **95% CI** | ***p*** |  | ***B*** | **95% CI** | ***p*** |  | ***B*** | **95% CI** | ***p*** |
| Model 1^3^ | |  |  |  |  |  |  |  |  |  |  |  |  |  |  |  |
|  | Former smoker  *[ref never smoker]* | 0.11 | 0.07; 0.14 | <0.001 |  | 0.10 | 0.06; 0.13 | <0.001 |  | 0.19 | 0.15; 0.22 | <0.001 |  | 0.13 | 0.10; 0.17 | <0.001 |
|  | Current smoker  *[ref never smoker]* | 0.33 | 0.27; 0.38 | <0.001 |  | 0.31 | 0.26; 0.37 | <0.001 |  | 0.54 | 0.48; 0.60 | <0.001 |  | 0.37 | 0.31; 0.42 | <0.001 |
|  | Current smoker  *[ref former smoker]* | 0.22 | 0.16; 0.28 | <0.001 |  | 0.22 | 0.16; 0.28 | <0.001 |  | 0.36 | 0.29; 0.42 | <0.001 |  | 0.23 | 0.17; 0.29 | <0.001 |
| Model 2^4^ | |  |  |  |  |  |  |  |  |  |  |  |  |  |  |  |
|  | Former smoker  *[ref never smoker]* | 0.08 | 0.05; 0.11 | <0.001 |  | 0.04 | 0.004; 0.07 | 0.029 |  | 0.13 | 0.10; 0.17 | <0.001 |  | 0.05 | 0.01; 0.08 | 0.010 |
|  | Current smoker  *[ref never smoker]* | 0.22 | 0.17; 0.28 | <0.001 |  | 0.22 | 0.17; 0.28 | <0.001 |  | 0.40 | 0.34; 0.45 | <0.001 |  | 0.27 | 0.22; 0.33 | <0.001 |
|  | Current smoker  *[ref former smoker]* | 0.14 | 0.09; 0.20 | <0.001 |  | 0.19 | 0.13; 0.25 | <0.001 |  | 0.26 | 0.20; 0.32 | <0.001 |  | 0.23 | 0.17; 0.29 | <0.001 |
| Model 3^5^ | |  |  |  |  |  |  |  |  |  |  |  |  |  |  |  |
|  | Former smoker  *[ref never smoker]* | 0.08 | 0.05; 0.11 | <0.001 |  | 0.04 | 0.01; 0.07 | 0.022 |  | 0.13 | 0.10; 0.17 | <0.001 |  | 0.05 | 0.02; 0.09 | 0.003 |
|  | Current smoker  *[ref never smoker]* | 0.22 | 0.17; 0.28 | <0.001 |  | 0.23 | 0.17; 0.28 | <0.001 |  | 0.40 | 0.34; 0.45 | <0.001 |  | 0.28 | 0.22; 0.33 | <0.001 |
|  | Current smoker  *[ref former smoker]* | 0.14 | 0.09; 0.20 | <0.001 |  | 0.19 | 0.13; 0.25 | <0.001 |  | 0.26 | 0.20; 0.32 | <0.001 |  | 0.22 | 0.16; 0.29 | <0.001 |
| ^1^ Possible range 1-6.  ^2^ Score of 5 or 6.  ^3^ Unadjusted.  ^4^ Adjusted for age, gender, ethnicity, income, and key worker status.  ^5^ Adjusted for age, gender, ethnicity, income, key worker status, and chronic physical health conditions. | | | | | | | | | | | | | | | | |

| **Supplementary Table 3.** Associations between smoking status and uncertainty and unwillingness to vaccinate against Covid-19: unweighted data | | | | | | | | |
| --- | --- | --- | --- | --- | --- | --- | --- | --- |
|  | | **Undecided** | | |  | **Unwilling** | | |
| **Descriptive data** | | **%** | **95% CI** | **-** |  | **%** | **95% CI** | **-** |
|  | Never smoker | 20.0 | 19.4; 20.5 | - |  | 9.6 | 9.2; 10.0 | - |
|  | Former smoker | 18.2 | 17.3; 19.1 | - |  | 11.2 | 10.4; 11.9 | - |
|  | Current smoker | 25.9 | 24.1; 27.8 | - |  | 16.6 | 15.1; 18.2 | - |
| **Multinomial logistic regressions** | | **RR** | **95% CI** | ***p*** |  | **RR** | **95% CI** | ***p*** |
| Model 1^1^ | |  |  |  |  |  |  |  |
|  | Former smoker  *[ref never smoker]* | 0.91 | 0.85; 0.98 | 0.008 |  | 1.16 | 1.06; 1.26 | 0.001 |
|  | Current smoker  *[ref never smoker]* | 1.59 | 1.44; 1.77 | <0.001 |  | 2.12 | 1.87; 2.40 | <0.001 |
|  | Current smoker  *[ref former smoker]* | 1.75 | 1.56; 1.96 | <0.001 |  | 1.83 | 1.60; 2.10 | <0.001 |
| Model 2^2^ | |  |  |  |  |  |  |  |
|  | Former smoker  *[ref never smoker]* | 0.92 | 0.86; 0.99 | 0.023 |  | 1.14 | 1.05; 1.25 | 0.004 |
|  | Current smoker  *[ref never smoker]* | 1.36 | 1.23; 1.52 | <0.001 |  | 1.69 | 1.49; 1.92 | <0.001 |
|  | Current smoker  *[ref former smoker]* | 1.48 | 1.32; 1.66 | <0.001 |  | 1.48 | 1.29; 1.70 | <0.001 |
| Model 3^3^ | |  |  |  |  |  |  |  |
|  | Former smoker  *[ref never smoker]* | 0.94 | 0.87; 1.01 | 0.084 |  | 1.18 | 1.07; 1.29 | <0.001 |
|  | Current smoker  *[ref never smoker]* | 1.39 | 1.25; 1.54 | <0.001 |  | 1.73 | 1.52; 1.97 | <0.001 |
|  | Current smoker  *[ref former smoker]* | 1.48 | 1.31; 1.66 | <0.001 |  | 1.47 | 1.28; 1.70 | <0.001 |
| ^1^ Unadjusted.  ^2^ Adjusted for age, gender, ethnicity, income, and key worker status.  ^3^ Adjusted for age, gender, ethnicity, income, key worker status, and chronic physical health conditions. | | | | | | | | |
